# An herbal drug combination identified by knowledge graph alleviates the clinical symptoms of plasma cell mastitis patients: a nonrandomized controlled trial

**DOI:** 10.1101/2022.12.01.22282958

**Authors:** Caigang Liu, Hong Yu, Guanglei Chen, Qichao Yang, Nan Niu, Ling Han, Dongyu Zhao, Manji Wang, Yuanyuan Liu, Yongliang Yang

## Abstract

**Background:** Plasma cell mastitis (PCM) is a nonbacterial breast inflammation with severe and intense clinical manifestation yet treatment methods for PCM are still rather limited. Although the mechanism of PCM remains unclear, mounting evidences suggest that the dysregulation of immune system is closely associated with the pathogenesis of PCM. Drug combinations or combination therapy could exert improved efficacy and reduced toxicity through hitting multiple discrete cellular targets.

**Methods:** We have developed a knowledge graph architecture towards immunotherapy and systematic immunity that consists of herbal drug-target interactions with a novel scoring system to select drug combinations based on target-hitting rates and phenotype relativeness. To this end, we employed this knowledge graph to identify an herbal drug combination for PCM and we subsequently evaluated the efficacy of the herbal drug combination in clinical trial

**Results:** Our clinical data suggests that the herbal drug combination could significantly reduce the serum level of various inflammatory cytokines, downregulate serum IgA and IgG level, reduce the recurrence rate and reverse the clinical symptoms of PCM patients with improvements of general health status.

**Conclusions:** In summary, we reported that an herbal drug combination identified by knowledge graph can alleviate the clinical symptoms of plasma cell mastitis patients. We demonstrated that the herbal drug combination holds great promise as an effective remedy for PCM, acting through the regulation of immunoinflammatory pathways and improvement of systematic immune level. In particular, the herbal drug combination could significantly reduce the recurrence rate of PCM, a major obstacle for PCM treatment. Our data suggests that the herbal drug combination is expected to feature prominently in future PCM treatment.

**Funding:** C. Liu’s lab was supported by grants from the Public Health Science and Technology Project of Shenyang (Grant: 22-321-32-18); Y. Yang’s laboratory was supported by the National Natural Science Foundation of China (Grant: 81874301), the Fundamental Research Funds for Central University (Grant: DUT22YG122) and the Key Research project of ‘be Recruited and be in Command’ in Liaoning Province (Personal Target Discovery for Metabolic Diseases).

**Clinical trial number:** ClinicalTrials.gov: NCT05530226

## Introduction

Plasma cell mastitis (PCM) represents as a serious inflammatory condition of breast that occurs in young and middle-aged females at non-pregnant and non-lactating period^1^. The main histopathological characteristics of PCM is the infiltration of plasma cells and lymphocytes in breast tissue^2^. Interestingly, PCM shares similarity with breast cancer in the perspective of macroscopical or microscopical characteristics^3^. On the other hand, mounting evidences suggest that the dysregulation of immune system is closely associated with the pathogenesis of PCM. Recently, the incidence rate of PCM is quickly rising yet the treatment methods for PCM are still rather limited. In clinical practice, surgical resection and hormone therapy remains as two major treatments for PCM. Unfortunately, neither surgical resection nor hormone therapy could prevent recurrence of PCM. Moreover, the serious side effects for hormone therapy are still problematic^4^. Therefore, the discovery of effective PCM treatment or therapeutics with minimal side effects is clearly warranted.

Traditional Chinese Medicine (TCM) has a rather long history for the prevention and treatment of complex diseases in eastern Asia^5-7^. Moreover, for decades, TCM has been often used as alternative or complementary medicine in the west. Indeed, Chinese herbal compounds has been successfully applied in the treatment of plasma cell mastitis (PCM) in conjunction with western medicine^8^. In clinical practice, TCM refers to herbal entity prescription or formulae (also called as ‘Fangji’) which may exhibit coordinating or synergistic effects through the combination of multiple herb drugs^9^. However, the design of formulae in TCM is solely based on the principle of ‘syndrome differentiation’ according to the medicinal properties of herbal entities. Moreover, the molecular mechanisms for the ‘formulae’ in TCM remains rather elusive.

Knowledge graph has emerged as an advanced technology in the field of artificial intelligence which is able to connect entities in a graph based on their existing intricate relationships^10^. In particular, knowledge graph can enable the rational design and identification of combination therapies for a specific disease or phenotypes^11^. Recently, we developed and constructed a knowledge graph for the discovery of herbal drug combination towards immunotherapy and systematic immunity. Subsequently, we identified a synergistic combination of herbal drugs for PCM via a scoring system based on target hitting rates and phenotype relativeness. To verify our concept of design, we conducted a clinical trial experiment for the drug combination of herbal compounds mentioned above (ClinicalTrials.gov Registration: NCT05530226). Strikingly, our clinical results demonstrated that the herbal drug combination identified by knowledge graph can markedly suppress various inflammatory cytokines in serum, restore clinical symptoms and reduce the recurrence rates of PCM patients with improved global health status.

## Results and Discussion

In previous study, we collected and compiled 240 targets for immunotherapy and systematic immunity from literature data^12^. Recently, we collected 345 entities of herbal drugs documented in TCM books and herbal drugs announced by National Administration of Traditional Chinese Medicine through advanced text-mining techniques. The existing intricate relationships between the herbal drugs and immunotherapy targets were also extracted and compiled via advanced text-mining techniques and manual curation for the construction of knowledge graph. We defined an ontology list consisting of 13 ontology terms describing the relations (edges) between herbal drug entities and the immunotherapy targets based on manual curation of literature data (**Supplemental Materials, Edge_ontology_terms**). Moreover, we collected the attributes of the medicinal properties for each herbal compound from Pharmacopoeia of China^13^. Totally, we compiled and integrated 64 attributes of the medicinal properties for herbal drug entities into the knowledge graph (**Supplemental Materials**, attributes of the medicinal properties). These medicinal properties are useful throughout the design of herbal drug combination. Finally, we built the knowledge graph via Neo4j and Py2Neo tools which consists of 895 nodes and 2197 edges (**Figure 1** and **Figure supplement 1**), which can be visited online (http://www.ikgg.org/).

**Figure 1.**
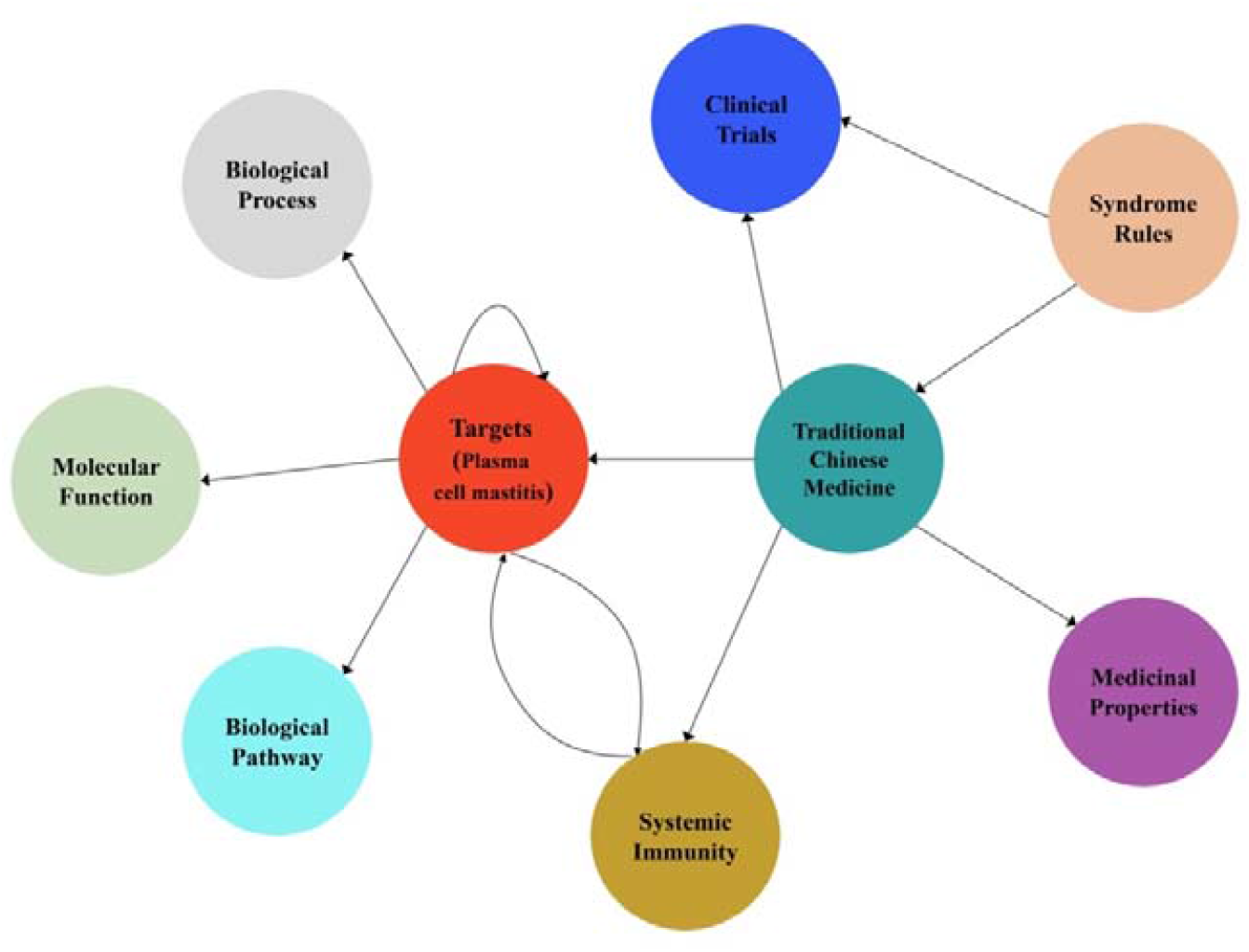
Schematic diagram of the knowledge graph for the drug discovery of plasma cell mastitis in the present work.

Subsequently, we employed a scoring system (or so-called recommendation system) to asses and predict synergistic herbal drug combination from the knowledge graph. Of note, the scoring function is able to identify those herbal drug combinations that are most related with specific phenotypes as well as herbal drug combinations that are able to hit most discrete cellular targets, yet still following the principle of ‘syndrome differentiation’ as described in Pharmacopoeia of China (**Materials and Methods**). To this end, we used this scoring function to select herbal drug combinations consisting of eight herbal entities. We chose to identify drug combinations with eight entities because ‘formulae’ consisting of eight drugs are regarded as ‘essence combination’ in TCM community. In short, we employed a combination generator that is able to randomly generate drug combinations with eight herbal drug entities for ten rounds, each of which consists of 1,000 random drug combinations. All the generated drug combinations from the ten rounds were further ranked and evaluated. Noteworthy, the scoring results of the ten rounds presented as normal distributions (**Figure 2**). The top twenty combinations from each round ranked by the scoring function were further curated and inspected by experts in TCM. Remarkably, we identified a specific drug combination which was ranked among top twenty choices in all ten rounds of calculation. The combination consists of eight herbal drug entities including ‘Fructus forsythiae’, ‘Herba violae’, ‘Uniflower swisscentaury root’, ‘Danshen’, ‘Astragalus’, ‘Taraxacum’, ‘Liquorice’ and ‘Honeysuckle’ (see **Figure 2**).

**Figure 2.**
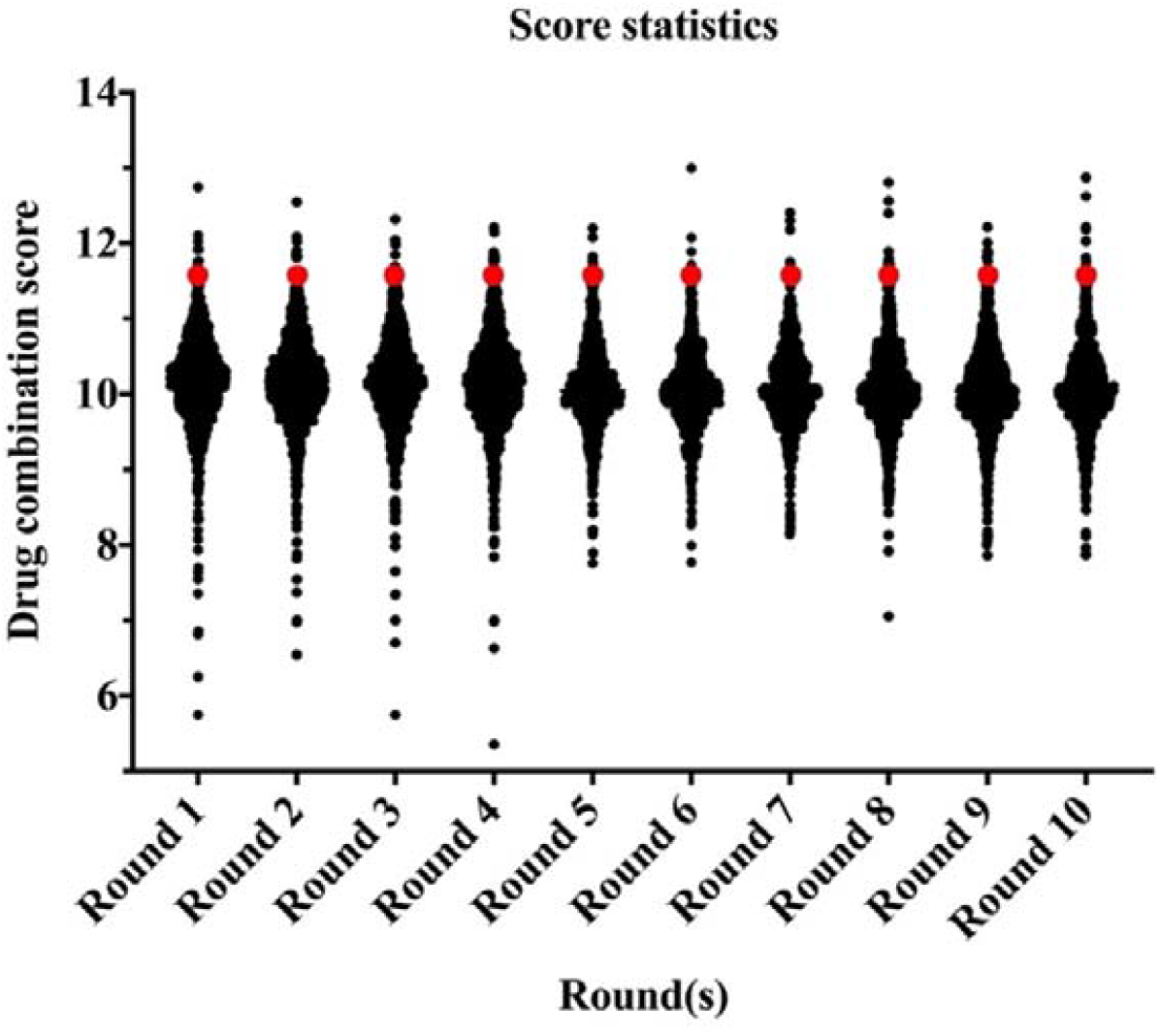
The score statistics of ten rounds of random herbal drug combinations (1,000 random combination from each round) for the treatment of plasma cell mastitis. Noteworthy, the scoring results of the ten rounds presented as normal distributions. The herbal drug combination identified for the clinical trial was among the top twenty choices in all the ten rounds and marked in red dots.

Next, we extracted the subgraph for the herbal drug combination mentioned above and created a network diagram for the drug combination via Cytoscape tools^14^ (**Figure 3**). In total, the eight herbal drug entities in the combination regulate 46 cellular targets related to immunotherapy and systematic immunity such as HIF-1, iNOS, IL-17, IL-6, IL-1β, mTOR, NLPR3, PD-L1, STAT3, TGF-β, TLR2 and TLR4 etc. (**Figure 3**). Noteworthy, the medicinal properties of the eight drug entities could be classified into three major categories of ‘Heat-clearing and detoxicating’, ‘Qi-tonifying’ and ‘Blood-activating menstruation regulating’. Moreover, we conducted pathway analysis for the herbal drug combination for plasma cell mastitis. Interestingly, we revealed that the herbal drug combination may modulate a few pathways related to systematic immunity including ‘Toll-like receptors cascades’, ‘MAP kinase activation’, Adaptive immune system’, ‘Growth hormone receptor signaling’, ‘Cytokine signaling in immune system’ and ‘Innate immune system’ via reactome knowledgebase^15^ (**Figure 4 and Supplemental Materials**). We believe all these may account for the therapeutic profiles of the herbal drug combination towards PCM.

**Figure 3.**
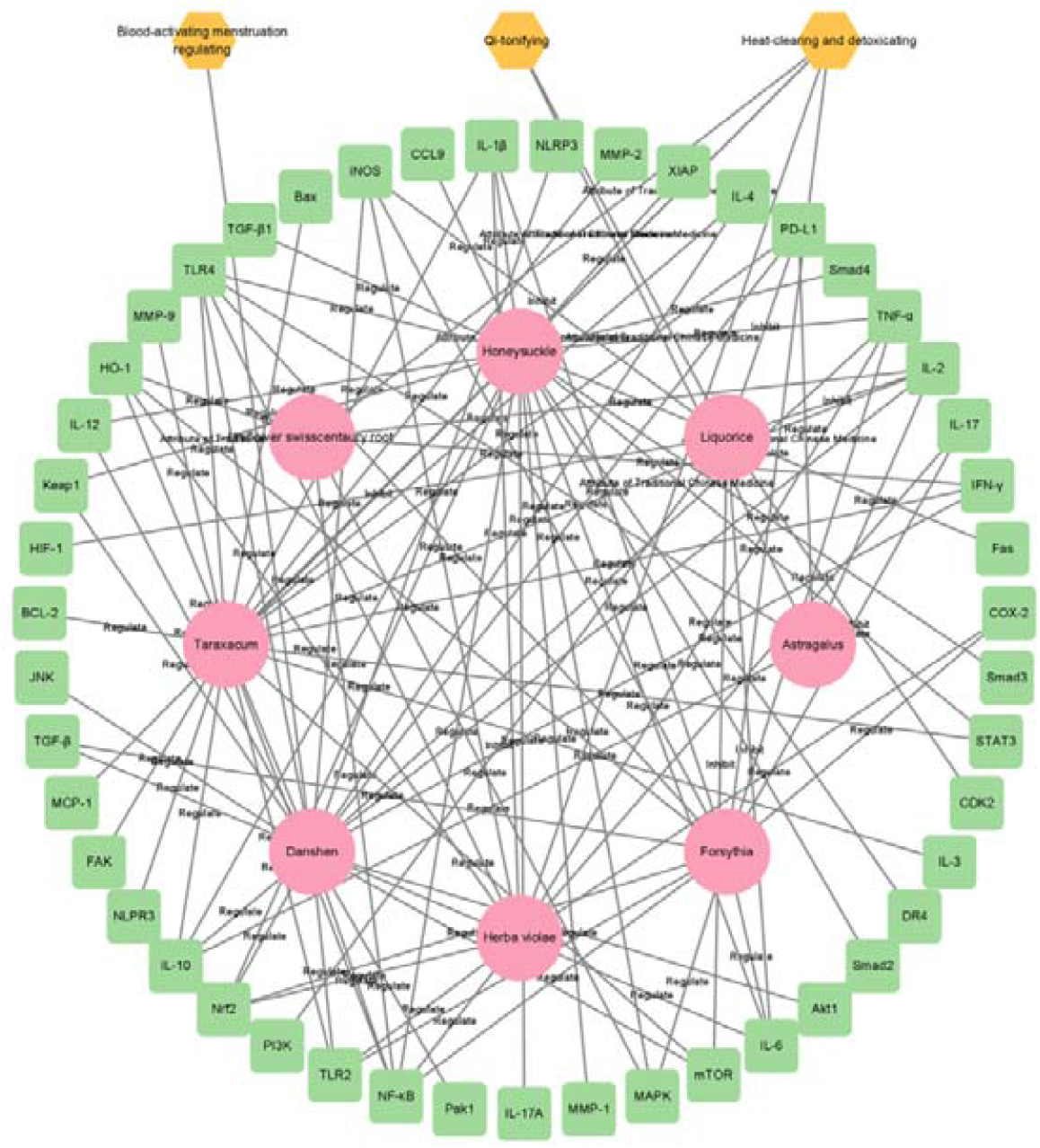
Network diagram of the herbal drug combination consisting of eight entities including ‘Honeysuckle’, ‘Taraxacum’, ‘Astragalus’, ‘Danshen’, ‘Forsythia’, ‘Herba violae’, ‘Liquorice’ and ‘Uniflower swisscentaury root’ displayed in red circles. In total, 46 cellular targets related to systemic immunity were hit by the drug combination such as NLPR3, IL-17, TLR4, STAT3, IL-6, iNOS and TLR2 et al which were displayed in green box model. Three major medicinal attributes (properties) were identified for these herbal drug entities including ‘Heat-clearing and detoxicating’, ‘Qi-tonifying’ and ‘Blood-activating menstruation regulation’.

**Figure 4.**
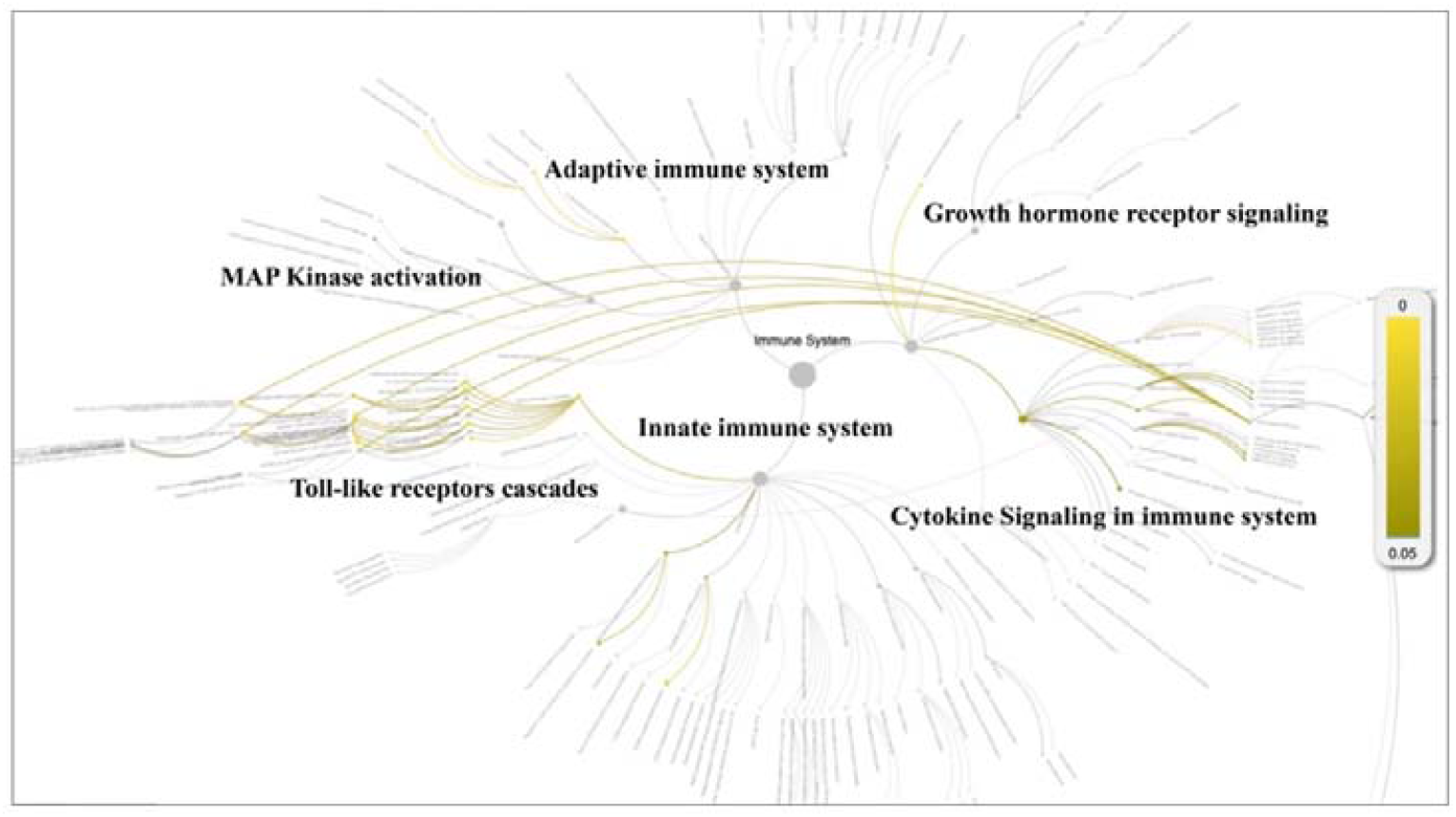
Pathway analysis for the potential cellular targets of the herbal drug combination via reactome knowledgebase. The statistically significant pathways were highlighted and displayed in yellow color (P<0.05).

Subsequently, we want to evaluate the efficacy of herbal drug combination in clinical trial for PCM patients (Clinicaltrials.gov number: NCT05530226). The ‘Jun-Chen-Zuo-Shi’ principle was examined for the herbal drug combination (‘Formulae’) by TCM experts and the dosage for each drug entity from the drug combination was adjusted by TCM experts. The clinical trial is a unrandomized, open label single arm study investigating the efficacy and safety of the herbal drug combination. To reveal the therapeutic effects of TCM drug combination, we selected patients who were treated with western medicine in the real world as comparison. Therefore, the two groups of patients were divided into TCM treatment group (experimental group) and western medicine treatment group (control group). All patients were diagnosed with PCM by biopsy of breast tissue before recruited into the clinical trial. Patients in the CG group (the control group) were orally treated with methylprednisolone whereas the patients in the EG group (the experimental or treatment group) were orally treated with herbal drug combination (**Materials and Methods**). Of note, methylprednisolone is a standard corticosteroid for the treatment of inflammatory conditions in clinical practice^16^ and therefore methylprednisolone is used in the CG group.

Efficacy was assessed every 2 cycles and the results were summarized after six months of treatment. The baseline characteristics are shown in **Table S1**. Strikingly, our results demonstrated that a few inflammatory cytokines in the serum including IL-2, IL-4, IL-6, IFN-γ, IL-1β and TNF-α were significantly downregulated in PCM patients after treatment of herbal drug combination as compared to the CG group treated with methylprednisolone (**Figure 5**). We chose to measure these cytokines in the experiments because they are often regarded as serum cytokine markers during the pathogenic development of PCM^17^. In addition, we found that serum level of IgA and IgG level were markedly suppressed in the treatment group of herbal drug combination as compared to the control group (**Figure 6**). Of note, both IgA and IgG have been found to be crucial diagnostic serum markers for PCM patients^18^. Moreover, IgA is regarded as a major component of mucosal immunity which is closely related to the pathogenesis of PCM^19,20^. Therefore, our data suggests that the herbal drug combination may enable the regulation of mucosal immunity and consequently downregulate IgA and IgG serum level. Furthermore, we conducted the standard Quality of Life questionnaire studies for PCM patients in the clinical experiment. Our results implicated that symptom score, pain score and global health status of PCM patients are significantly improved after treatment of the herbal drug combination as compared to the control group (**Figure 7**). Noteworthy, our results demonstrated that the recurrence rate of PCM patients in the treatment group were reduced to 3.75% as compared to the recurrence rate of 12.5% in the control group (**Table 1**). Moreover, the incidence rate of adverse events of PCM patients in the treatment group were reduced to 6.25% as compared to the recurrence rate of 11.25% in the control group (**Table 1**). In addition, we observed that the clinical symptoms of PCM patients in the EG group such as swelling, abscess and fistula were reversed (**Figure 8, Table_S2 in Supplemental File**) after treatment of herbal drug combinations. The clinical symptom score in the EG group is ∼4.68 as compared to the clinical symptom score of ∼5.98 in the CG group (**Table 2**). These results suggest that the herbal drug combination may achieve better efficacy for the treatment of PCM as compared to methylprednisolone.

**Table 1.** Comparison of operation rate, recurrence rate and incidence of adverse reactions between the two groups (EG: experimental group; CG: control group).

| **Groups** | **N**<br>**(Patients)** | **Operation** | **Recurrence** | **Incidence of**<br>**adverse events** |
| --- | --- | --- | --- | --- |
| **EG** | 80 | 25 (31.25%) | 3 (3.75%) | 5 (6.25%) |
| **CG** | 80 | 47 (58.75%) | 10 (12.5%) | 9 (11.25%) |
| **X<sup>2</sup>** |  | 12.22 | 4.103 | 1.252 |
| **p** |  | <0.001 | 0.043 | 0.263 |

**Table 2.** Clinical symptom scores between the two groups in the clinical trial (EG: experimental group; CG: control group; The clinical symptom rating scale for PCM was displayed in Table_S2 in the Supplemental File).

| **Groups** | **N**<br>**(Patients)** | **Baseline** | **Treated** | **t** | **p** |
| --- | --- | --- | --- | --- | --- |
| **EG** | 80 | 13.90±2.37 | 4.68±3.40 | 22.19 | <0.001 |
| **CG** | 80 | 13.423±2.70 | 5.98±3.68 | 14.282 | <0.001 |
| **t** |  | 1.189 | 2.323 |  |  |
| **p** |  | 0.236 | 0.021 |  |  |

**Figure 5.**
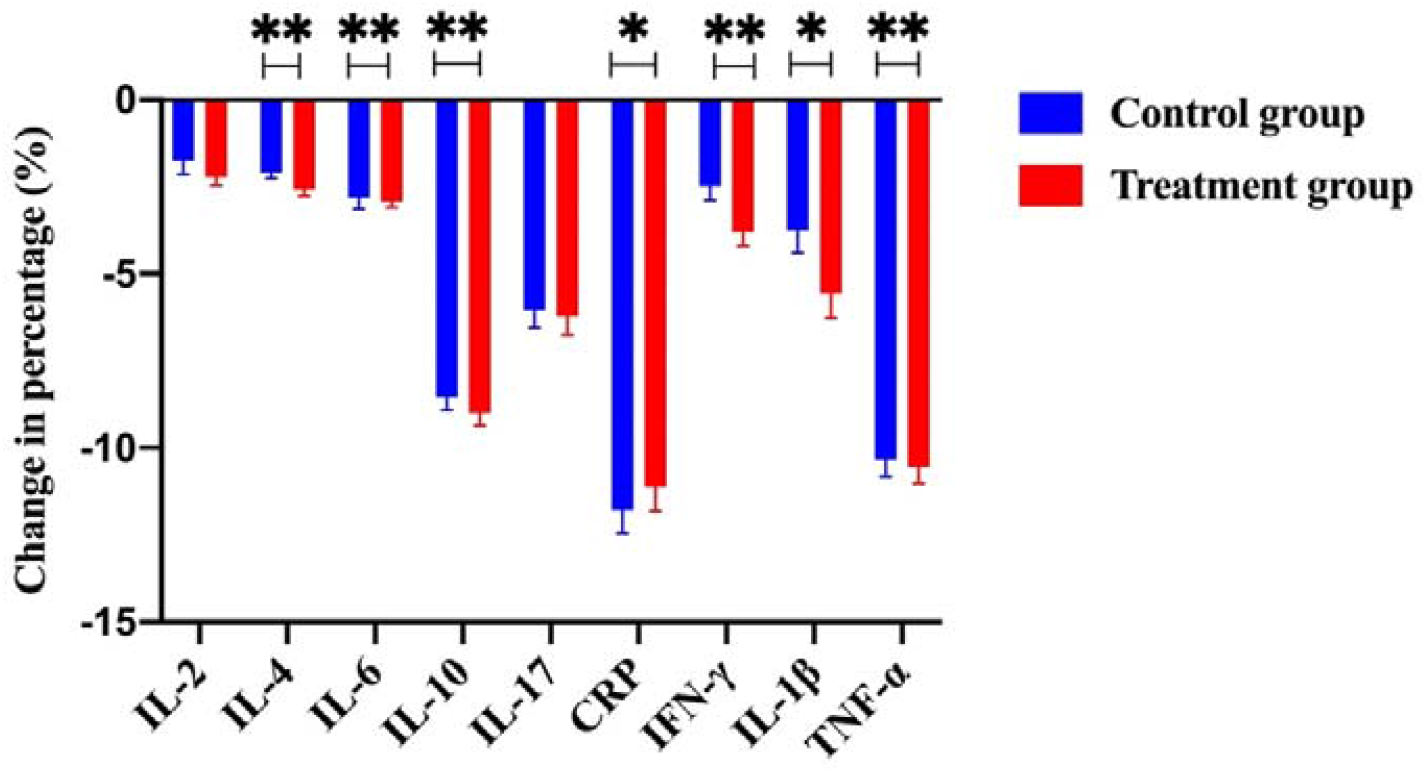
The change of percentage for numerous immunological cytokines including IL-2, IL-4, IL-6, IL-10, IL-17, CRP, IFN-γ, IL-1β and TNF-α from the control group and the treatment group (with TCM treatment). Notably, a few key cytokines such as IL-2, IL-4, IFN-γ, IL-1β and TNF-α were significantly downregulated in the treatment group as compared to the control group. **Figure 5 – source data**. Patients’ raw data of serum cytokines from the control group (without TCM treatment) and the treatment group (with TCM treatment) in the clinical trial (ClinicalTrials.gov: NCT05530226).

**Figure 6.**
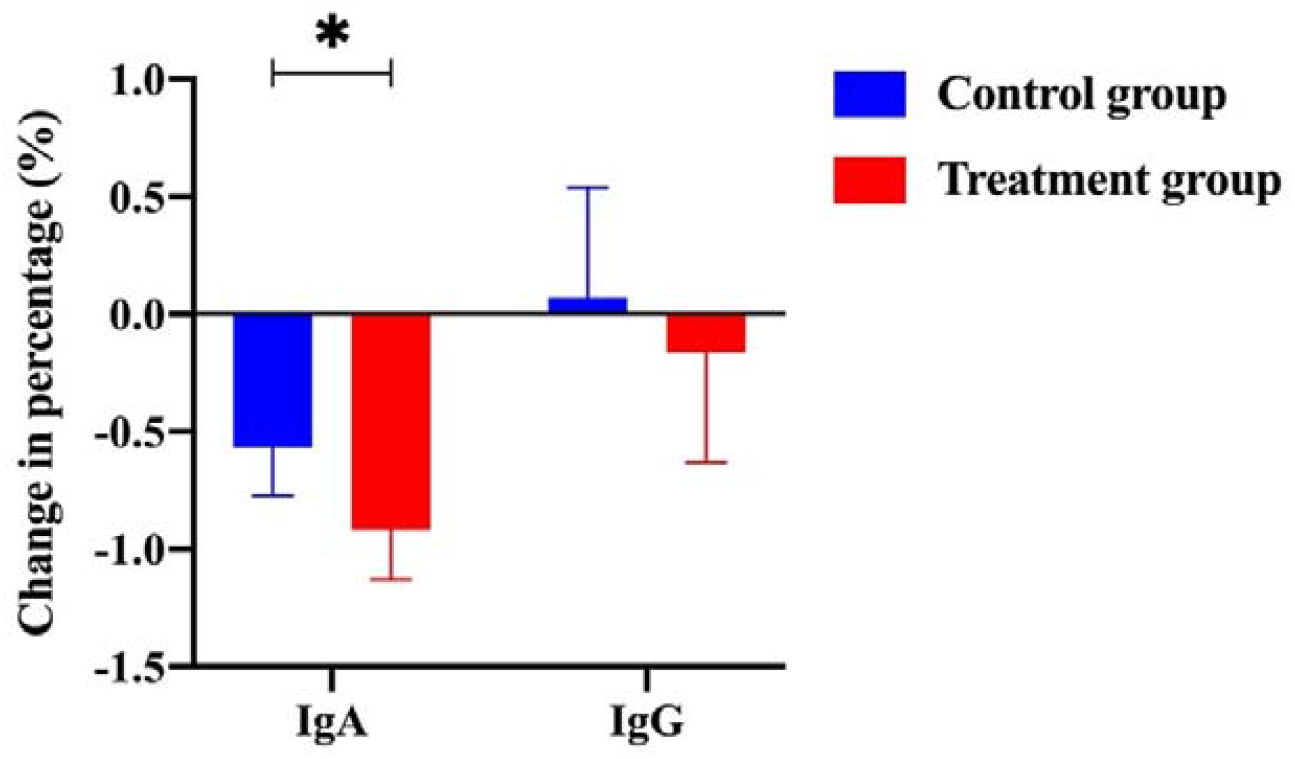
The change of percentage for IgA level and IgG level from the control group and the treatment group (with TCM treatment). **Figure 6 – source data**. Patients’ raw data of serum IgA and IgG level from the control group (without TCM treatment) and the treatment group (with TCM treatment) in the clinical trial (ClinicalTrials.gov: NCT05530226).

**Figure 7.**
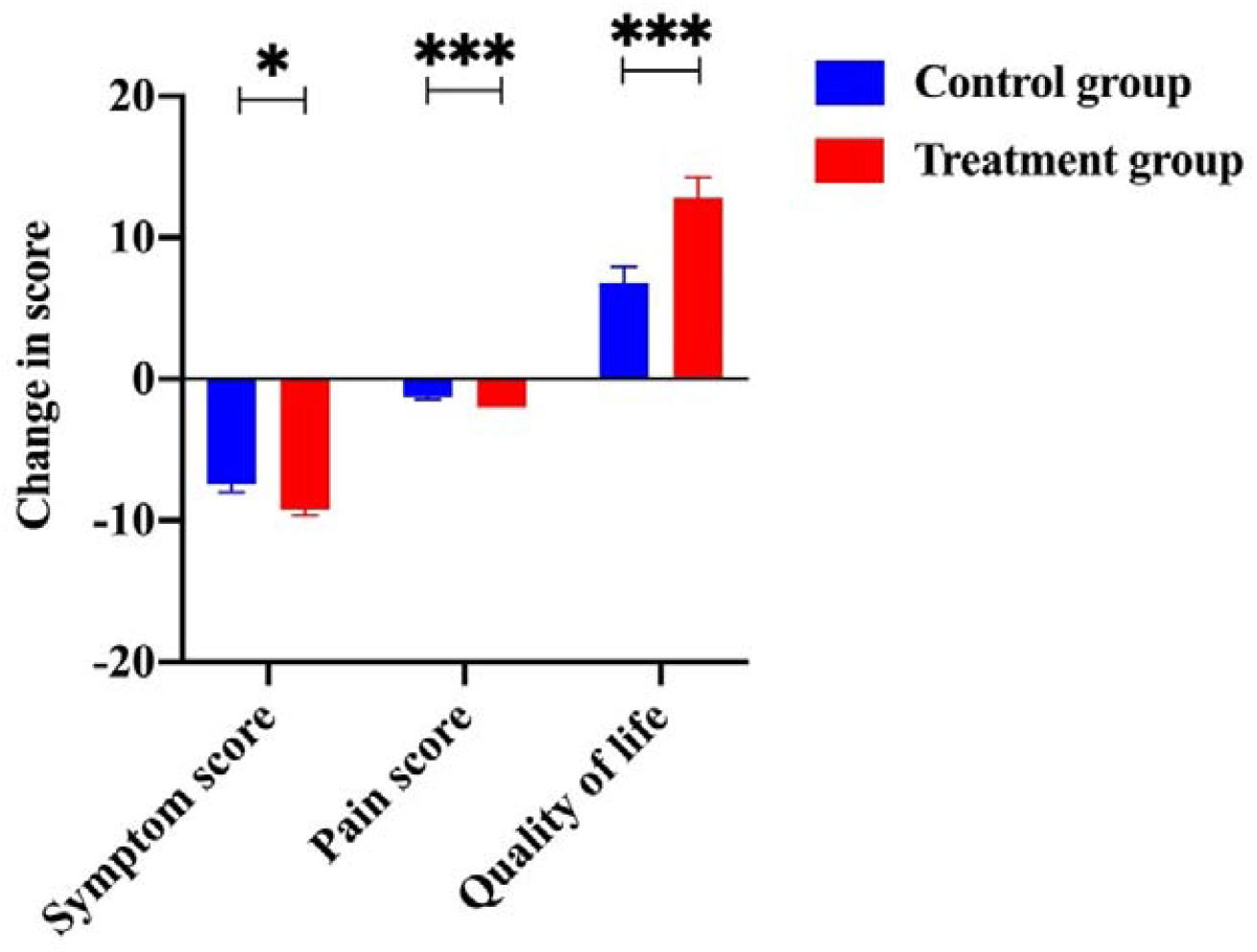
The change of scores from items including symptom, pain and living quality. The scores of symptoms, pain and living quality from the treatment group were significantly improved as compared to the control group. **Figure 7 – source data 1**. Patients’ raw data of symptom scores from the control group (without TCM treatment) and the treatment group (with TCM treatment) in the clinical trial (ClinicalTrials.gov: NCT05530226). **Figure 7 – source data 2**. Patients’ raw data of pain scores from the control group (without TCM treatment) and the treatment group (with TCM treatment) in the clinical trial (ClinicalTrials.gov: NCT05530226). **Figure 7 – source data 3**. Patients’ raw data of quality of life from the control group (without TCM treatment) and the treatment group (with TCM treatment) in the clinical trial (ClinicalTrials.gov: NCT05530226).

**Figure 8.**
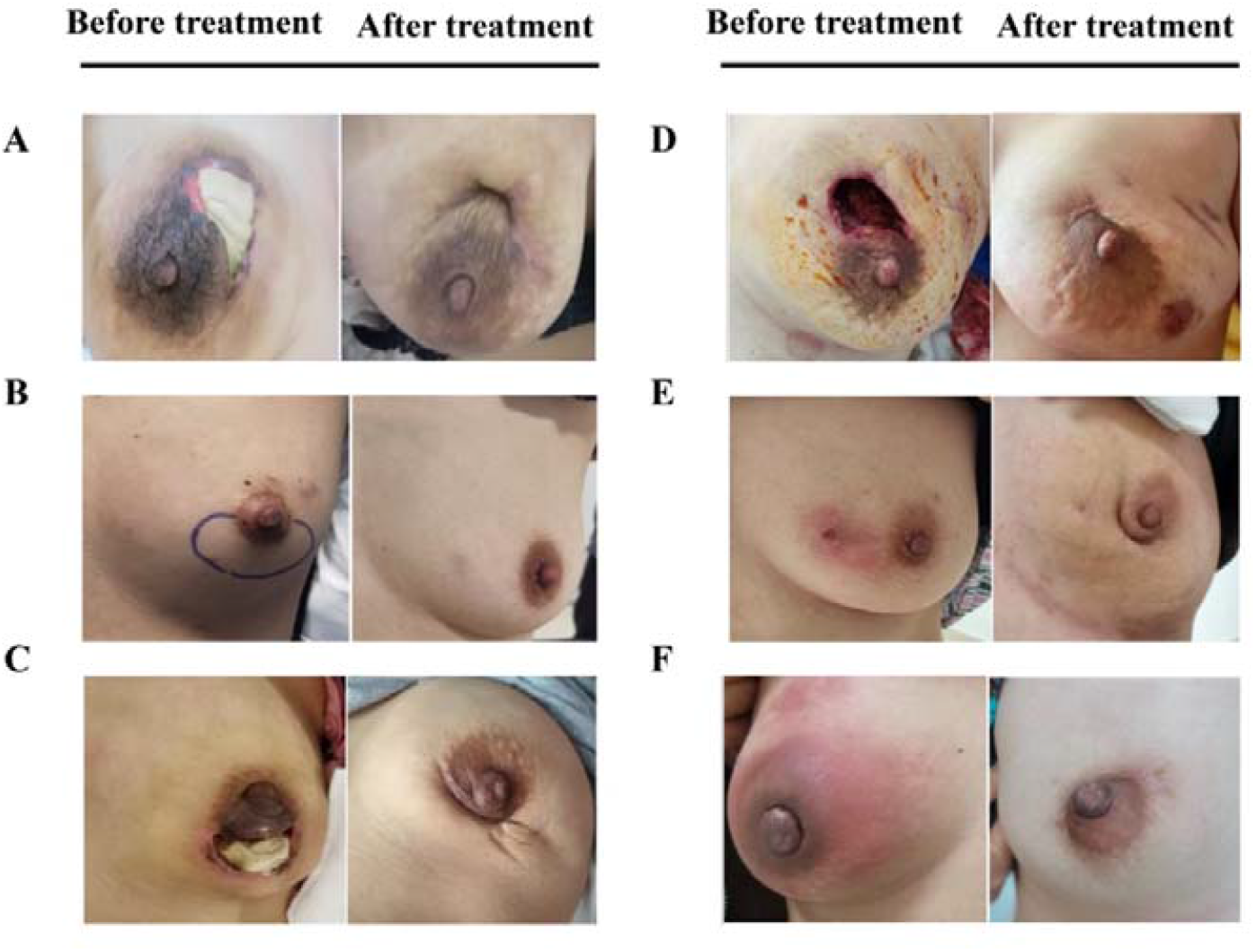
The comparison of whole breast for six representative PCM patients in the EG group before treatment and after treatment (written informed consents have been provided from all the patients).

With the increasing amount of biomedical data, the traditional drug discovery campaign has been revolutionized with the aid of artificial intelligence techniques to accelerate the process and reduce the cost^21^. In recent years, knowledge graph, a technique that can provide structured relations among entities and unstructured semantic relations associated with entities, has been introduced into the domain of drug discovery^11^. Although the pathogenesis of PCM remains largely unclear, there have been numerous reports implicating that the overactivation of immunoinflammatory pathways play an important role in the development of PCM^22^. The major advantage of using Traditional Chinese Medicine is that herbal drug combination can hit multiple discrete targets related to immunoinflammatory pathways with improved efficacy and reduced toxicity. Herein, for the first time, we showcase an example that identifies an herbal drug combination via knowledge graph towards PCM. In contrast to using the conventional principle of ‘syndrome differentiation’, our knowledge graph consisting of intricate relations between herbal drug entities and immunotherapy targets coupled with scoring functions are able to automatically identify novel herbal drug combinations which can hit most discrete targets, making this strategy unique in the TCM community. Although we acknowledge that the inclusion of chemical ingredients from the herbal drugs may impact the outcome of our analysis and design, unfortunately, the inclusion of chemical ingredients in the knowledge graph is rather technically difficult due to the limited and incomplete datasets for the herbal drugs in the field of TCM. Nevertheless, our strategy captures the prominent feature of design for drug combinations towards a complex disease such as PCM. In the future, we plan to include multiple types of omics data such as genomic, transcriptomic, proteomic, metagenomic or metabolomics data into the knowledge graph to reveal novel targets and enable novel drug discovery.

In the present study, our results revealed that the herbal drug combination identified by knowledge graph could suppress a few key immunoinflammatory cytokines, enhance the systematic immune levels and significantly reduce the recurrence rates of PCM patients. Of note, recurrence has become one major obstacle after surgical resection for PCM treatment in clinical practice. On the other hand, hormone therapy may increase the risk of side effects for PCM patients. Therefore, our approach of herbal drug combination may provide a new avenue for PCM treatment with less recurrence rate and reduced incidence rate of adverse events.

## Conclusion

In summary, we report the identification and clinical assessment of an herbal drug combination towards Plasma cell mastitis (PCM). We demonstrated that the herbal drug combination holds great promise as an effective remedy for PCM, acting through the regulation of immunoinflammatory pathways and improvement of systematic immune level. In particular, the herbal drug combination could significantly reduce the recurrence rate of PCM, a major obstacle for PCM treatment. Our data suggests that the herbal drug combination is expected to feature prominently in the future PCM treatment. Moreover, these promising results underscore the potential of knowledge graph to identify drug combinations or other novel therapeutics across multiple types of human disorders.

## Data Availability

All data refereed to in the manuscript are available to the public.

## Acknowledgments

Y. Yang’s laboratory was supported by the National Natural Science Foundation of China (Grant: 81874301), the Fundamental Research Funds for Central University (Grant: DUT22YG122) and the Key Research project of ‘be Recruited and be in Command’ in Liaoning Province (Personal Target Discovery for Metabolic Diseases); C. Liu’s lab was supported by grants from the National Natural Science Foundation of China (No. 81572609), China Medical University Major Construction Project (No. 2017ZDZX05) and Liaoning Colleges Innovative Talent Support Program (Cancer Stem Cell Origin and Biological Behavior).

## Author Contributions

Q.Y., and G.C. performed data mining and data curation and constructed the knowledge graph; Q.Y., Y.L. and Y.Y. designed the scoring system for the knowledge graph; Q.Y., and Y.Y. participated in the design and styling of the web interface; D.Z. helped to collect and analyze the medicinal properties of the herbal drug entities; L.H. and N.N. participated in the design of herbal drug combination product; M.W. and G.C. conducted analysis of clinical data and clinical pictures of the PCM patients; H.Y. and N.N. coordinated the clinical trial experiments and registration; C. L. and Y.Y. provided funding for the project; Y.Y. and C.L. initiated, coordinated the whole project; Y.Y. wrote the manuscript.

## Declaration of Ethnics

The protocol was approved by the Institutional Review Board (IRB) of the China Medical University (approval number: 2021PS024T). This study was registered with ClinicalTrials.gov: NCT05530226. All patients provided written informed consent.

## Materials and Methods

### Construction of knowledge graph towards immunotherapy

We employed data mining techniques to collect and compile 240 targets of immunotherapy and systematic immunity from PubMed database. Next, we collected and compiled 345 herbal drug entities officially released by the National Health Commission of China and National Administration of Traditional Chinese Medicine. The intricate relations between the herbal drug entities and the immunotherapy targets were extracted from the PubMed database. These intricate relations were subjected to further manual curation. We used thirteen ontology terms (see **Supplemental Materials**) to describe the intricate relations (edges) in the knowledge graph. Moreover, 64 attributes of the medicinal properties for the herbal drug entities were collected and compiled from Pharmacopoeia of China. Finally, we built the knowledge graph via Neo4j and Py2Neo tools which consists of 895 nodes and 2197 edges.

### Scoring system of the knowledge graph

To this end, we developed a scoring system to asses and predict synergistic drug combination of herbal drug entities (number of drugs, n) as below,

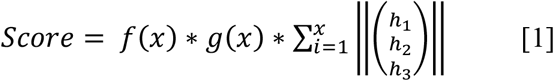

Herein, *f*(*x*) represents as the penalty function. *f*(*x*) value will be set to 0 if the medicinal properties of the drug combination fall into the contraindication rules in Pharmacopoeia of China. Otherwise, *f*(*x*) value will be set to 1.

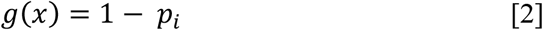

*g*(*x*) is the target diversity function as above and *p*_*i*_ is calculated as *p*_*i*_ = *w*/*t*; *t* refers to the total number of targets within the knowledge graph, *w* refers to the total number of overlapping targets that the drug combination may hit. Hence, the target diversity function can be used as a measure to assess the diversity of the targets that the drug combination may hit. In another word, if each drug entity in the combination hits distinct targets, the *g*(*x*) value will be set to 1. The last term of the scoring system is used as a measure to assess the relativeness of each drug entity in the combination and calculated as follows,

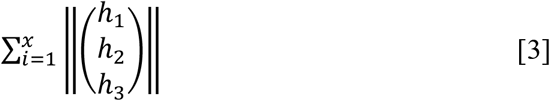

In brief, *h*_1_ represents the target hitting rates of each drug entity in the combination and was calculated as follows, *h*_1_ = *n*_*i*_/*t*; *n*_*i*_ is the number of hitting targets for each drug entity in the combination; Again, *t* refers to the total number of targets constituting the knowledge graph for the disease; Noteworthy, the concept of hitting rates towards discrete targets has been used in the scoring function for the selection of synergistic drug combinations^23^. *h*_2_ represents the phenotype relativeness of each drug entity in the combination and *h*_2_ = *c*_2_ * 1/*x*, where *x* is the number of drug entities in the combination and *c*_2_ is the parameter; Namely, if the drug entity is related to the phenotype of the disease (co-occurrence with the disease phenotype in the literature), then *c*_2_ value is set to 1 otherwise *c*_2_ value is set to 0; *h*_3_ represents the literature relativeness or confidence of each drug entity in the combination and calculated as follows,

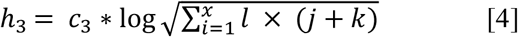

in which *l* is the number of studies/publications that validated the association of drug entity with the specific disease (herein in the knowledge graph refers to cancer immunotherapy), *j* and *k* refer to if the relations of the drug entity with the disease have been validated in cell lines or patient (or animal) tissues, respectively. Namely, if the drug entity was validated in cancer cell lines or patient tissues, the *j* or *k* value will be set to 1, respectively. Otherwise, the *j* or *k* value will be set to 0; *c*_3_ is the parameter and set to 1 here. Therefore, herein, a high score of *h*_3_ implicates that the drug combination is more relevant to cancer immunotherapy with high confidence of literature relativeness. Collectively, our scoring system can be used to select those drug combinations that are most relevant with disease phenotypes and those drug combinations that are able to hit most discrete targets related to immunotherapy.

### Design of the clinical trial

In brief, 160 female patients diagnosed as plasma cell mastitis (PCM) in Shengjing Hospital Affiliated to China Medical University were recruited in the clinical trial between January 2021 to February 2022. Patients were randomly 1:1 divided into experimental group (EG) and control group (CG). Noteworthy, in order to demonstrate the therapeutic effect of TCM drug combination, we selected patients who were treated with western medicine in the real world during the same period. Therefore, the two groups of patients were divided into TCM treatment group (experimental group) and western medicine treatment group (control group). There was no significant difference in baseline data such as age, body mass index, clinical classification, marriage and child-bearing history between the two groups (**Supplemental File, Table_S1**). Patients in the CG group were orally treated with methylprednisolone tablets, 20mg/ day once a day. The patients in the EG group were orally treated with 20g/bag of herbal drug combination twice a day, once in the morning and once in the evening for 2 months. The herbal drug combination was prepared as granules in the following formulae: Taraxacum 15g, Fructus forsythiae 15g, Honeysuckle 10g, Uniflower swisscentaury root 8g, Herba violae 20g, Danshen 10g, Astragalus 20g, Liquorice 8g. The herbal drug combination in the form of granules was provided and prepared by Shengjing Hospital Affiliated to China Medical University.

### Clinical trial protocol

The clinical trial for the herbal drug combination was registered at ClinicalTrials.gov and entitled as “A Single Arm Study of Traditional Chinese Medicine for Plasma Cell Mastitis” with registration code of NCT05530226. The detailed clinical trial protocol has been provided a separate document in the Supplemental Files named as ‘PCM_Clinical_Protocol’.

### Measurement of serum inflammatory cytokines by ELSIA assay

Venous blood of the CG and EG groups were collected in sterile non-anticoagulant test tube before and after treatment. The immune transmission turbidimetry was used according to the procedure of CRP kit and automatic biochemical analyzer was used to detect the level of CRP. The levels of serum cytokines were measured by ELISA (Elabscience) according to the manufacturer’s instructions.

### Measurement of serum immunoglobulin level

The venous blood of PCM patients in the two groups were collected in sterile non-anticoagulant tube before and after treatment. The serum IgG and IgA were measured by rate scattering turbidimetry using Array 360 System automatic specific protein analyzer (Beckman Company, USA).

### Assessment of clinical symptoms of PCM patients

The clinical symptoms were evaluated by attending physician with board certification in pathology. The patients were scored before and after treatment according to the standard rating scale for PCM (see **Supplemental Materials**).

### Statistics

All data were evaluated as mean ± SEM. Statistical analysis of the quantitative multiple group comparisons was performed using the one-way analysis of variance (ANOVA) followed by Tukey’s test; whereas pairwise comparisons were performed using the t test by GraphPad Prism 8 (Graph Pad Software, La Jolla, CA, USA). Results were considered to be statistically significant with p<0.05.

## Figures

**Figure 1 - Figure supplement 1.**
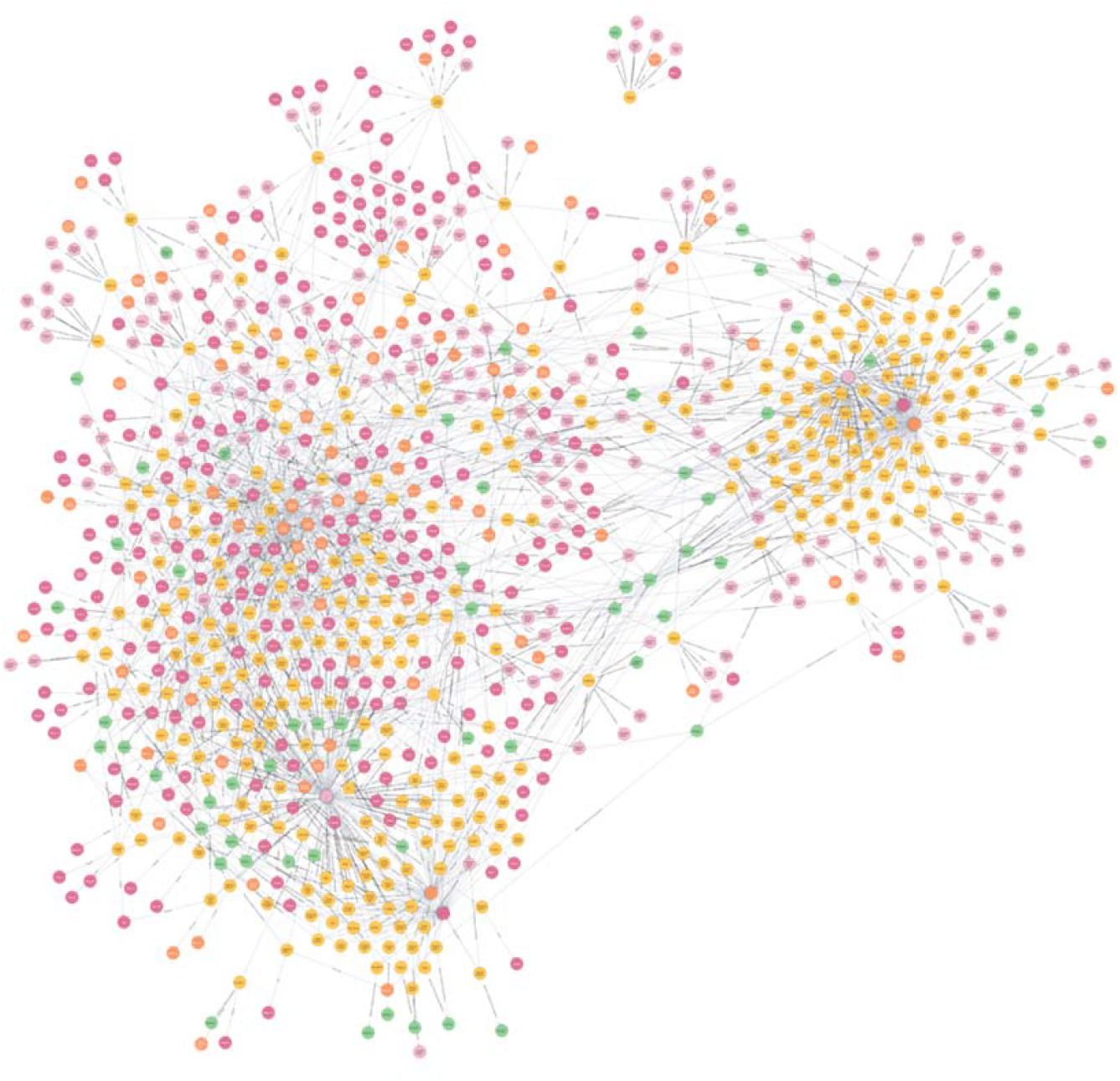
Snapshot of the medical knowledge graph towards immunotherapy. The knowledge graph was constructed by Neo4j and Python tools (http://www.ikgg.org/kghti).

## References

1. Cutler, M. Plasma-cell mastitis; report of a case with bilateral involvement. Br Med J 1, 94–96 (1949).

2. Yu, J.J., et al. Mouse model of plasma cell mastitis. J Transl Med 10 Suppl 1, S11 (2012).

3. Cheng, L., et al. Mastitis, a Radiographic, Clinical, and Histopathologic Review. Breast J 21, 403–409 (2015).

4. Fleming, L., et al. The impact of medication side effects on adherence and persistence to hormone therapy in breast cancer survivors: A quantitative systematic review. Breast 64, 63–84 (2022).

5. Tu, Y. The discovery of artemisinin (qinghaosu) and gifts from Chinese medicine. Nat Med 17, 1217–1220 (2011).

6. Tu, Y. Artemisinin-A Gift from Traditional Chinese Medicine to the World (Nobel Lecture). Angew Chem Int Ed Engl 55, 10210–10226 (2016).

7. Yin, X., et al. Effect of Electroacupuncture on Insomnia in Patients With Depression: A Randomized Clinical Trial. JAMA Netw Open 5, e2220563 (2022).

8. Zhang, J., Xu, J., Zhang, J. & Ren, Y. Chinese herbal compound combined with western medicine therapy in the treatment of plasma cell mastitis: A protocol for systematic review and meta-analysis. Medicine (Baltimore) 99, e22858 (2020).

9. Li, S., Zhang, B., Jiang, D., Wei, Y. & Zhang, N. Herb network construction and co-module analysis for uncovering the combination rule of traditional Chinese herbal formulae. BMC Bioinformatics 11 Suppl 11, S6 (2010).

10. Ye, Q., et al. A unified drug-target interaction prediction framework based on knowledge graph and recommendation system. Nat Commun 12, 6775 (2021).

11. Zeng, X., Tu, X., Liu, Y., Fu, X. & Su, Y. Toward better drug discovery with knowledge graph. Curr Opin Struct Biol 72, 114–126 (2022).

12. Zhang, Y., et al. Checkpoint therapeutic target database (CKTTD): the first comprehensive database for checkpoint targets and their modulators in cancer immunotherapy. J Immunother Cancer 8(2020).

13. Hao, Y.F. & Jiang, J.G. Origin and evolution of China Pharmacopoeia and its implication for traditional medicines. Mini Rev Med Chem 15, 595–603 (2015).

14. Reimand, J., et al. Pathway enrichment analysis and visualization of omics data using g:Profiler, GSEA, Cytoscape and EnrichmentMap. Nat Protoc 14, 482–517 (2019).

15. Jassal, B., et al. The reactome pathway knowledgebase. Nucleic Acids Res 48, D498–D503 (2020).

16. Lv, J., et al. Effect of Oral Methylprednisolone on Clinical Outcomes in Patients With IgA Nephropathy: The TESTING Randomized Clinical Trial. JAMA 318, 432–442 (2017).

17. Liu, Y., et al. Activation of the IL-6/JAK2/STAT3 pathway induces plasma cell mastitis in mice. Cytokine 110, 150–158 (2018).

18. Xing, M., Zhang, S., Zha, X. & Zhang, J. Current Understanding and Management of Plasma Cell Mastitis: Can We Benefit from What We Know? Breast Care (Basel) 17, 321–329 (2022).

19. Betts, C.B., et al. Mucosal Immunity in the Female Murine Mammary Gland. J Immunol 201, 734–746 (2018).

20. Bharathan, M. & Mullarky, I.K. Targeting mucosal immunity in the battle to develop a mastitis vaccine. J Mammary Gland Biol Neoplasia 16, 409–419 (2011).

21. Yang, Y., Adelstein, S.J. & Kassis, A.I. Target discovery from data mining approaches. Drug Discov Today 14, 147–154 (2009).

22. Liu, Y., et al. IL-6/STAT3 signaling pathway is activated in plasma cell mastitis. Int J Clin Exp Pathol 8, 12541–12548 (2015).

23. Jin, W., et al. Deep learning identifies synergistic drug combinations for treating COVID-19. Proc Natl Acad Sci U S A 118(2021).

